# Changes in cardiorespiratory fitness and body mass index due to COVID-19 mitigation measures in Austrian children aged 7 to 10 years

**DOI:** 10.1101/2021.04.09.21255185

**Authors:** Gerald Jarnig, Johannes Jaunig, Mireille N.M. van Poppel

**Affiliations:** Institute of Human Movement Science, Sport and Health, University of Graz, Graz, Austria

## Abstract

**Importance:** Previous studies showed reduced self-reported physical activity levels in children due to the coronavirus disease 2019 (COVID-19) mitigation measures, whereas limited data is available for objectively assessed health parameters.

**Objective:** To examine the influence of these measures on the longitudinal development of cardiorespiratory fitness (CRF) and body mass index (BMI) of primary school children.

**Design:** Cohort study with baseline measurements in September 2019, before the COVID-19 mitigation measures and follow-ups in June and September 2020.

**Setting:** Twelve randomly selected primary schools in urban and rural districts of Klagenfurt, Austria.

**Participants:** Legal guardians of 860 children provided written consent. A total of 764 children (88.8%) aged 7–10 years completed all measurements and were included for analyses.

**Exposure:** COVID-19 mitigation measures.

**Main Outcomes and Measures:** The study was planned as a randomized controlled trial, but analyzed as a longitudinal study due to stopped intervention because of COVID-19 mitigation regulations. CRF was measured with a 6-min run test. Height and weight were objectively measured. Age- and gender-specific national and international standard deviation scores (SDS) were calculated for CRF and BMI. Changes over time were analysed using ANOVAs. Secondary analyses were performed for subgroups divided by gender and sports club membership.

**Results:** From September 2019 to September 2020, CRF SDS decreased by −1.06 (95% CI, - 1.13 to −1.00), with a similar rate of decrease in both boys and girls. In June 2020, BMI SDS had increased by 0.12 (95% CI, 0.06-0.16), and in September 2020 by 0.16 (95% CI, 0.12-0.20), compared to September 2019. The rate of increase in BMI SDS was higher in boys (0.23 [95% CI, 0.18-0.29]) than in girls (0.09 [95% CI, 0.04-0.15]). Over the 1-year period, the proportion of children with overweight or obesity increased from 20.3% to 24.1% (+3.8%, *P* < .001), according to International Obesity Taskforce thresholds.

**Conclusions and Relevance:** COVID-19 mitigation measures have negative indirect consequences of on relevant health parameters of children. Since these mitigation measures continued after our last assessments, consequences will have increased. Collaborative efforts are required to negate and reverse these effects on children’s health, to prevent long-term negative health consequences.

**KEY POINTS:** *Question:* Do the COVID-19 mitigation measures have an impact on cardiorespiratory fitness (CRF) and body mass index (BMI) of primary school children?

*Findings:* In this longitudinal study, which included 764 primary school children, substantial reductions were found in CRF, which is a parameter related to long-term cardiovascular risk factors. In addition, increases in BMI standard deviation scores and in the proportion of children who are overweight or obese were evident.

*Meaning:* Collaborative efforts are required to negate and reverse these effects on children’s health, to prevent long-term negative health consequences.

## INTRODUCTION

The indirect consequences of the coronavirus disease 2019 (COVID-19) pandemic are of great concern, especially the consequences for children. Various studies worldwide described the negative effects of pandemic mitigation measures on self- or proxy-reported levels of physical activity and sedentary behaviour of children and youth.^1-6^ The reported reduction in physical activity levels and the increase in sedentary behaviour will likely affect relevant health-related parameters, such as cardiorespiratory fitness (CRF), while indirectly affecting the body mass index (BMI).

CRF in childhood is an important health marker,^7^ and a higher CRF is related with lower levels of BMI, waist circumference, body fatness and a reduced prevalence of metabolic syndrome in later life.^8^ Childhood obesity is associated with increased cardiovascular risk factors^9^ and coronary heart disease.^10^ However, to our knowledge there are currently no studies on the effects of COVID-19 mitigation measures on objectively measured CRF and BMI in a representative sample of children.

Like in many other countries, children in Austria had very limited access to exercise and sport from March 2020 till September 2020, since playgrounds and sport facilities were closed in the spring of 2020, and they did not attend any physical education (PE) classes in school until September 2020 (Figure 1). Therefore, we aimed to examine the influence of COVID-19 mitigation measures on the longitudinal development of CRF and BMI of primary school children in Klagenfurt, Austria, from September 2019 to September 2020.

**Figure 1.**
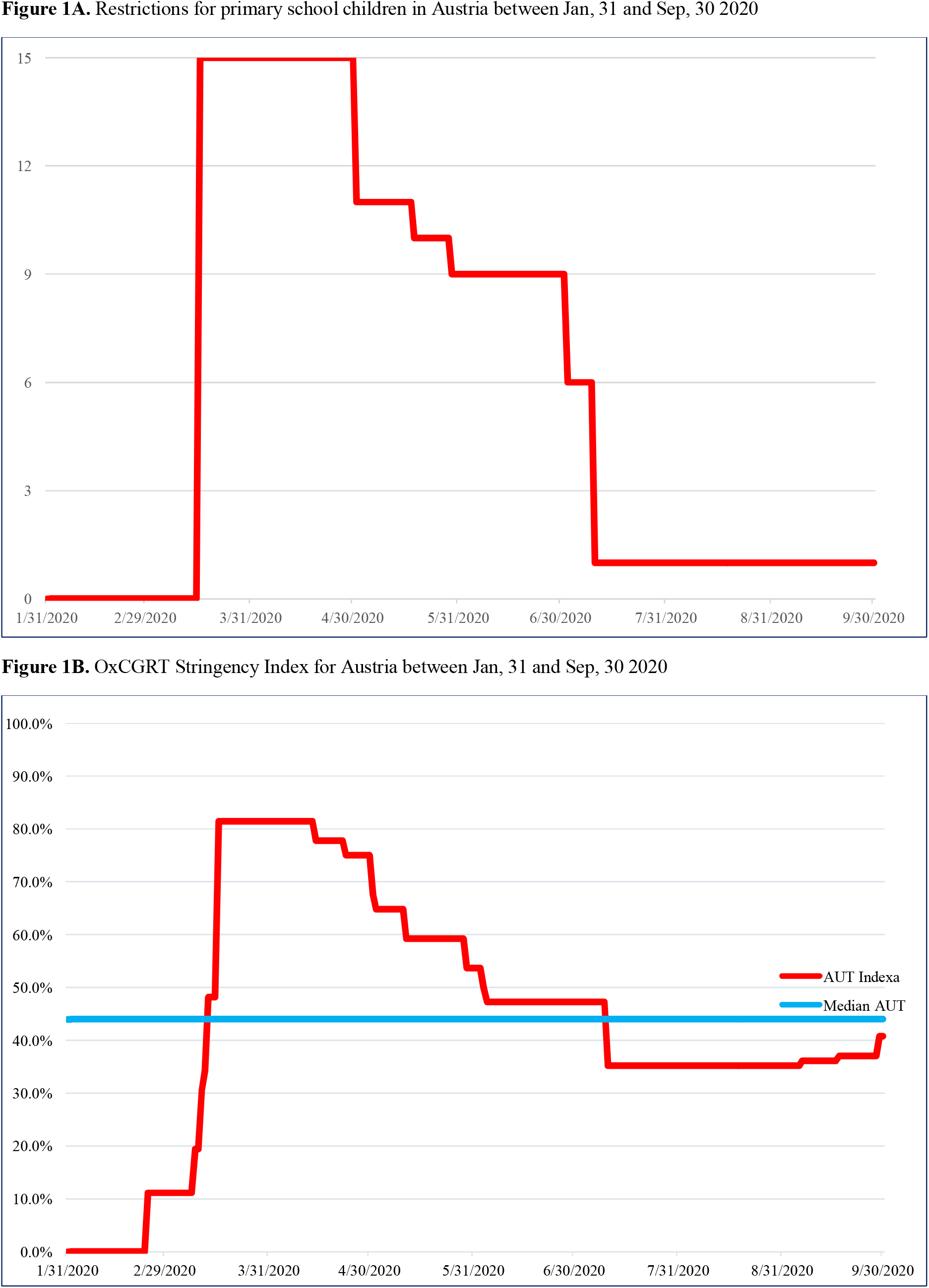
COVID-19 restrictions in Austria between Jan, 31 and Sep, 30 2020

## METHODS

This study was originally designed as a randomised controlled trial to evaluate the effects of a PE intervention on motor competence, CRF and health of primary school children. Due to COVID-19 regulations, the intervention had to be stopped in March 2020. We analysed the data as that of a longitudinal study and assessed the impact of COVID-19 mitigation measures on CRF and BMI of children aged 7 to 10 years. The study was registered in the German Clinical Trial Register (ID DRKS00023824) and approved by the Research Ethics Committee at the University of Graz, Styria, Austria (GZ. 39/23/63 ex 2018/19).

### Selection of schools and participants

A list of all 39 primary schools in both urban and rural districts of Klagenfurt, Austria, was used for the selection of schools. Using a random number generator, 12 schools were selected and randomised into intervention and control groups, stratified by district. All schools consented to participate in the study. Inclusion criteria were as follows: age between 7–10 years at baseline and the physical ability to perform all motor competence tests in the test battery. Between May and June 2019, we invited all 1013 children attending these 12 schools to participate. Legal guardians of 860 (85%) children provided written consent for their children’s participation (Figure 2).

**Figure 2.**
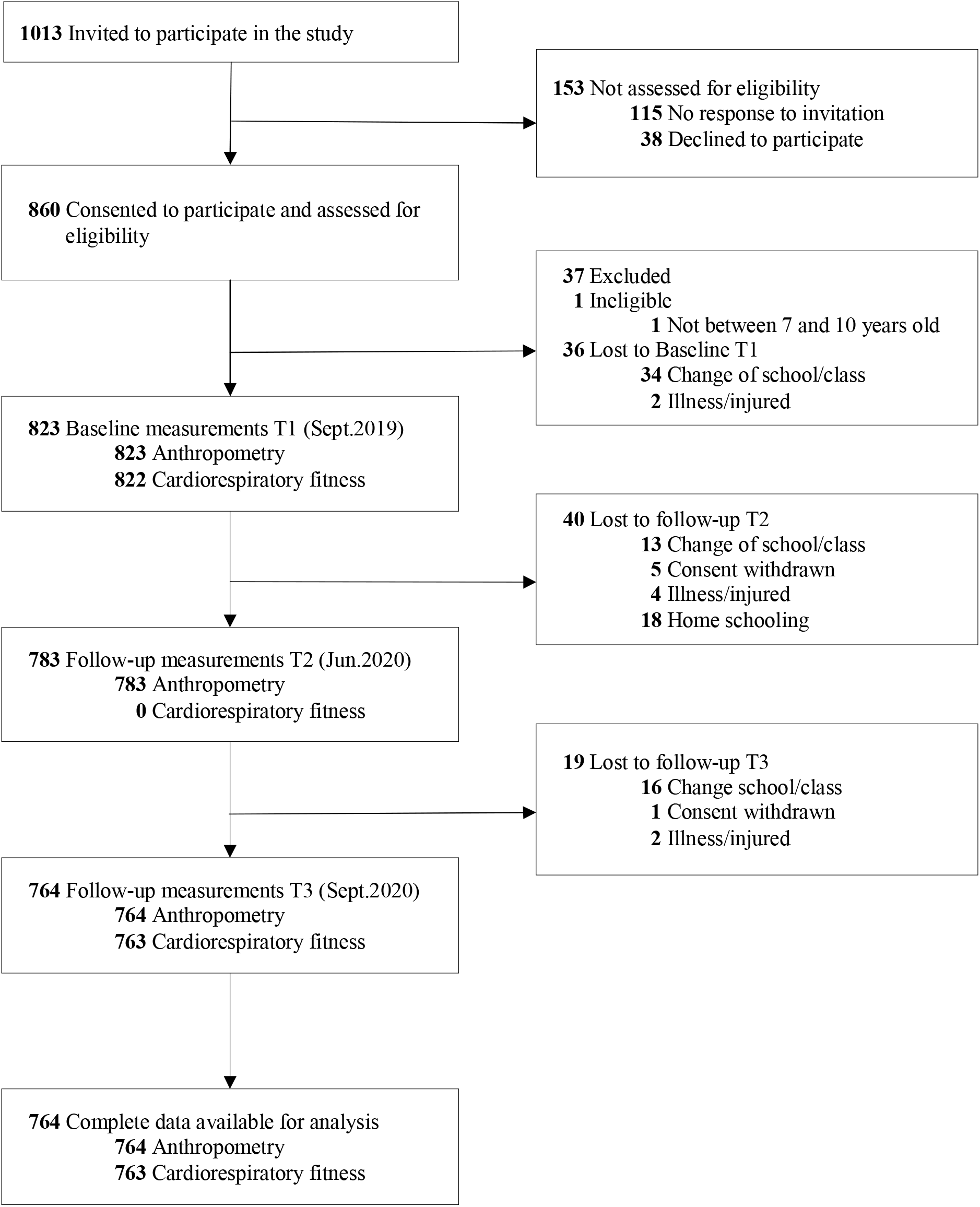
Flow diagram

### Procedures

Baseline measurements (T1) were performed in September and October 2019. When the lockdown restrictions were slowly relaxed (Figure 1B), the legal department of the Education Directorate of Carinthia allowed a second test phase (T2) in May and June 2020, under strict hygiene measures and regulations applicable at that time.^11^ The CRF test was not permitted at that time, since the minimum distance between participants could not be assured. In September and October 2020, the third test phase (T3) was conducted in which the complete test battery was performed.

The intervention started after baseline measurements, in October 2019. In the intervention group, external trainers and sports scientists planned and taught all PE classes. The intervention and usual PE classes (control group) were interrupted by the lockdown on 16 March 2020.^11^

The Oxford COVID-19 Government Response Tracker (OxCGRT) provides internationally comparable stringency levels for Austria at the study period (Figure 1B); a more detailed self-developed stringency level for the impact of the pandemic mitigation measures on children over time can be seen in Figure 1A. The precise description of this classification method, on the basis of the Austrian legislative decrees, accessible in the federal law gazette,^11^ is available in the eTable 1.

### Outcomes

In this study, the primary outcomes were the impact of the lockdown measures on CRF and BMI. The anthropometric data included height (cm) and weight (kg). Height was measured to the nearest 0.1 cm with a portable stadiometer (SECA 213, Hamburg, Germany). Weight was measured to the nearest 0.1 kg using an electronic weighing scale (BOSCH PPW4202/01, Nuremberg, Germany). The BMI was calculated as weight in kilograms divided by height in metres squared. As a measure of CRF, we used the 6-min endurance run test (6-min run),^12,13^ since this is the most relevant fitness parameter for future cardiovascular health status.^8^ The 6-min run was performed according to the protocol of the German Motor Test (GMT).^13^ Standardised deviation scores were calculated for both CRF and BMI, using age-and gender-specific national and international reference values. We used alternative reference values in sensitivity analyses to check for robustness of the findings. In addition, BMI was dichotomised, using various national and international reference thresholds for below and above the definition of overweight. Secondary analyses were performed for subgroups (divided by gender and by sports club membership), for changes in height (using reference values) and the examination of possible confounders (age, allocation to intervention, residential area (rural/urban), and school).

### Statistical analysis

Descriptive statistics were calculated for all three time points. Continuous variables are expressed as mean (M) and standard deviation (SD), and categorial variables as absolute values (n) and percentages (%). The analysed data only contains complete data for all measured time points (Figure 2), and no data imputation was performed.

#### Standardisation of anthropometric variables

Based on national and international age- and gender-specific references, standard deviation scores (SDS) for BMI and height were calculated. For national reference values, recent data from a large, representative Austrian sample were used.^14,15^ For comparison with international reference values, the International Obesity Taskforce (IOTF)^16^ and WHO^17^ references were considered. For both international references, calculations based on the LMS method^18^ were performed. The Austrian reference data were calculated using GAMLSS models (see Gleiss et al^15^ and Mayer et al^14^ for detailed statistical procedure). National reference values are expressed in BMI centile curves (ie, equicurves), representing BMI values corresponding to values at the age of 18 years (hereinafter referred to as EQUI BMI [AUT]). The IOTF reference uses the same centile curve approach^16^, but it is expressed in SDS (referred to as BMI SDS [IOTF]). This approach allows BMI thresholds be on a continuum to those used for adults (ie, overweight BMI ≥ 25 kg/m^2^). WHO reference values are expressed in SDS as well (referred to as BMI SDS [WHO]), but it uses +1 SDS as a threshold for overweight.^17^ Therefore, the dichotomous classification results (underweight and normal weight vs. overweight and obesity) of the three references were calculated using their given thresholds, respectively. For height SDS values, only AUT and WHO references were available.

#### Standardisation of the 6-min run test

To compare the results of the 6-min run test (raw score) with different existing reference values, z-scores were created based on the most recent age- and gender-specific references values. Since no reference values are available for Austrian children at this age, German references were applied. The most recent percentile tables from the Düsseldorfer Modell (DüMo; collected 2011–2018)^12^ are available for the use of LMS method, to compute z-scores (referred to as 6-min run SDS [DüMo]). The reference values from the GMT^13^ uses the traditional z-score standardisation (referred to as 6-min run SDS [GMT]).

#### Changes over time

In multilevel mixed models, with individuals and school as the two random levels, possible clustering of outcomes within schools and the influence of possible confounders (age, intervention, residential area) were assessed. Since we observed no significant clustering and influence of possible confounders, further analyses were performed using three-way analysis of variance with repeated measurements (ANOVAs). Gender and sports club membership (yes/no) entered the models as between subject effects, and the measurement time points (three for BMI and two for 6-min run test) were entered as within subject effects. In case of non-sphericity, a correction according to Greenhouse–Geisser was carried out. Homogeneity was tested with the Harley’s F-max test. To visualise interaction effects, the scores in the figures were plotted by gender and by sports club in separate lines/bars, with statistics for significant interaction effects described within. To account for over-proportional increase in height SDS, multivariate ANOVAs (ie, MANOVAs) were used to combine standardised height and BMI values.

Differences in the dichotomised BMI classification (underweight and normal weight vs. overweight and obese) were analysed for the total sample and for the subgroups (gender by sports club membership) using the Cochrane’s Q test for a possible time effect. The Dunn test was subsequently used to investigate the effects between the pairwise measurement time points (T1–T2, T1–T3 and T2–T3).

All tests were two-sided, with a p-value less than 0.05 considered statistically significant. Alpha level correction for post-hoc tests were performed using Bonferroni correction. In the case of ANOVAs and MANOVAs, partial eta squared (η_p_^2^) was used to determine the size of the effect (≥ 0.01 = small, ≥ 0.06 = medium, ≥ 0.14 = large effect), hence only an at least small effect (η_p_^2^ ≥ 0.01) was considered relevant. All statistical calculations were performed using SPSS Version 26 (IBM Corp. Released 2019. IBM SPSS Statistics for Windows, Armonk, NY: IBM Corp).

## RESULTS

In September 2019, 823 children participated in the baseline measurements. Fifty-nine (60 for CRF; both 7%) children did not complete the assessments in all three measurements time points, and they were excluded from analyses, resulting in 764 children with complete data on anthropometry and 763 on CRF (Figure 2). The cluster school (n=12) showed no significant influence as a random factor in multilevel mixed model analyses for all primary outcomes. No confounding and no significant interaction effects were found for possible confounders (age, intervention and residential area; data not shown), which were not included in further analyses. Sample characteristics for the total sample as well as for boys and girls separately are shown in Table 1 (eTable 2).

**Table 1.** Sample characteristics for the total sample as well as for boys and girls separately

|  |  | Sep 2019 | Jun 2020 | Sep 2020 |
| --- | --- | --- | --- | --- |
| **Age, mean (SD), y** |  | 8.26 (0.69) | 8.96 (0.69) | 9.24 (0.69) |
| **Girls, No. (%)** |  | 383 (50.1%) | 383 (50.1%) | 383 (50.1%) |
| **Member of sports club, No. (%)** |  | 322 (42.1%) | 322 (42.1%) | 322 (42.1%) |
| **Rural region, No. (%)** |  | 451 (59.0%) | 451 (59.0%) | 451 (59.0%) |
| **6-min run, mean (SD), m** | total | 917.0 (141.0) | ND | 815.0 (134.3) |
|  | girls | 871.2 (121.4) | ND | 777.4 (118.7) |
|  | boys | 963.1 (144.5) | ND | 852.8 (138.4) |
| **6-min run SDS (DüMo), mean (SD)** | total | 0.49 (1.13) | ND | -0.57 (0.97) |
|  | girls | 0.40 (1.08) | ND | -0.64 (0.96) |
|  | boys | 0.58 (1.17) | ND | -0.50 (0.98) |
| **Weight, mean (SD), kg** | total | 29.9 (7.2) | 33.1 (8.5) | 34.5 (8.9) |
|  | girls | 29.9 (7.1) | 33.1 (8.3) | 34.5 (8.9) |
|  | boys | 29.8 (7.3) | 33.1 (8.6) | 34.4 (9.0) |
| **Height, mean (SD), cm** | total | 132.2 (6.6) | 136.4 (6.9) | 138.1 (6.9) |
|  | girls | 131.7 (6.8) | 136.0 (7.1) | 137.8 (7.1) |
|  | boys | 132.7 (6.4) | 136.8 (6.6) | 138.4 (6.6) |
| **BMI SDS (IOTF), mean (SD)** | total | 0.37 (1.08) | 0.49 (1.10) | 0.53 (1.10) |
|  | girls | 0.45 (1.08) | 0.54 (1.10) | 0.56 (1.13) |
|  | boys | 0.28 (1.08) | 0.45 (1.10) | 0.50 (1.07) |

Sample size for anthropometric data, N = 764; for 6-min run test, N = 763.
BMI = body mass index, cm = centimeter, DüMo = 6-min run SDS based on the Düsseldorfer Modell (Stemper et al, 2020), IOTF = BMI SDS based on International Obesity Taskforce reference centile curves (Cole et al, 2012), kg = kilogram, m = meter, ND = not determined, SDS = standard deviation score.

**Table 2.** Children with overweight and obesity using IOTF, Austrian and WHO reference thresholds

|  |  | No. | Sep 2019<br>ow+ob | Jun 2020<br>ow+ob | Sep 2020<br>ow+ob |
| --- | --- | --- | --- | --- | --- |
|  |  |  | No. (%) | No. (%) | No. (%) |
| IOTF<br>(Cole et al) | total | 764 | 155 (20.3) | 169 (22.1) | 184 (24.1) |
|  | girls total | 383 | 91 (23.8) | 97 (25.3) | 103 (26.9) |
|  | girls sports club | 119 | 20 (16.8) | 24 (20.2) | 26 (21.8) |
|  | girls no sports club | 264 | 71 (26.9) | 73 (27.7) | 77 (29.2) |
|  | boys total | 381 | 64 (16.8) | 72 (18.9) | 81 (21.3) |
|  | boys sports club | 203 | 31 (15.3) | 34 (16.7) | 39 (19.2) |
|  | boys no sports club | 178 | 33 (18.5) | 38 (21.3) | 42 (23.6) |
| AUT<br>(Mayer et al) | total | 764 | 114 (14.9) | 132 (17.3) | 144 (18.8) |
|  | girls total | 383 | 57 (14.9) | 68 (17.8) | 70 (18.3) |
|  | girls sports club | 119 | 10 (8.4) | 13 (10.9) | 16 (13.4) |
|  | girls no sports club | 264 | 47 (17.8) | 55 (20.8) | 54 (20.5) |
|  | boys total | 381 | 57 (15.0) | 64 (16.8) | 74 (19.4) |
|  | boys sports club | 203 | 28 (13.8) | 31 (15.3) | 35 (17.2) |
|  | boys no sports club | 178 | 29 (16.3) | 33 (18.5) | 39 (21.9) |
| WHO<br>(de Onis et al) | total | 764 | 202 (26.4) | 235 (30.8) | 243 (31.8) |
|  | girls total | 383 | 112 (29.2) | 121 (31.6) | 119 (31.1) |
|  | girls sports club | 119 | 28 (23.5) | 30 (25.2) | 31 (26.1) |
|  | girls no sports club | 264 | 84 (31.8) | 91 (34.5) | 88 (33.3) |
|  | boys total | 381 | 90 (23.6) | 114 (29.9) | 124 (32.5) |
|  | boys sports club | 203 | 48 (23.6) | 59 (29.1) | 61 (30.0) |
|  | boys no sports club | 178 | 42 (23.6) | 55 (30.9) | 63 (35.4) |

AUT = Austrian reference values using centile curves for BMI 25.00 at the age of 18 as thresholds for overweight and above (Mayer et al, 2015), BMI = body mass index, IOTF = International Obesity Taskforce reference values using centile curves for BMI 25.00 at the age of 18 as thresholds for overweight and above (Cole et al, 2012), SDS = standard deviation score, ow+ob = overweight including obesity, WHO = World Health Organization reference values using +1 SDS percentiles as thresholds for overweight and above (de Onis et al, 2007).

### Change in CRF

A reduction in physical activity levels and sports participation will directly affect physical fitness, especially CRF. From September 2019 to September 2020, the mean distance the children were able to run in 6 minutes decreased from 917.0 (SD 141.1) to 815.0 (SD 134.3) metres. The mean 6-min run SDS (DüMo) decreased from 0.49 (SD 1.13) to −0.57 (SD 0.97) (η_p_^2^=0.554, *P*<.001), and for 6-min run SDS (GMT) from 0.15 (SD 1.09) to −0.96 (SD 1.01) (η_p_^2^=0.595, *P*<.001). Although children who were members of sports clubs had better CRF at all time points, the decrease in CRF over time was similar in all groups (Figure 3A). For detailed results of the ANOVAs see eTable 3.

**Figure 3.**
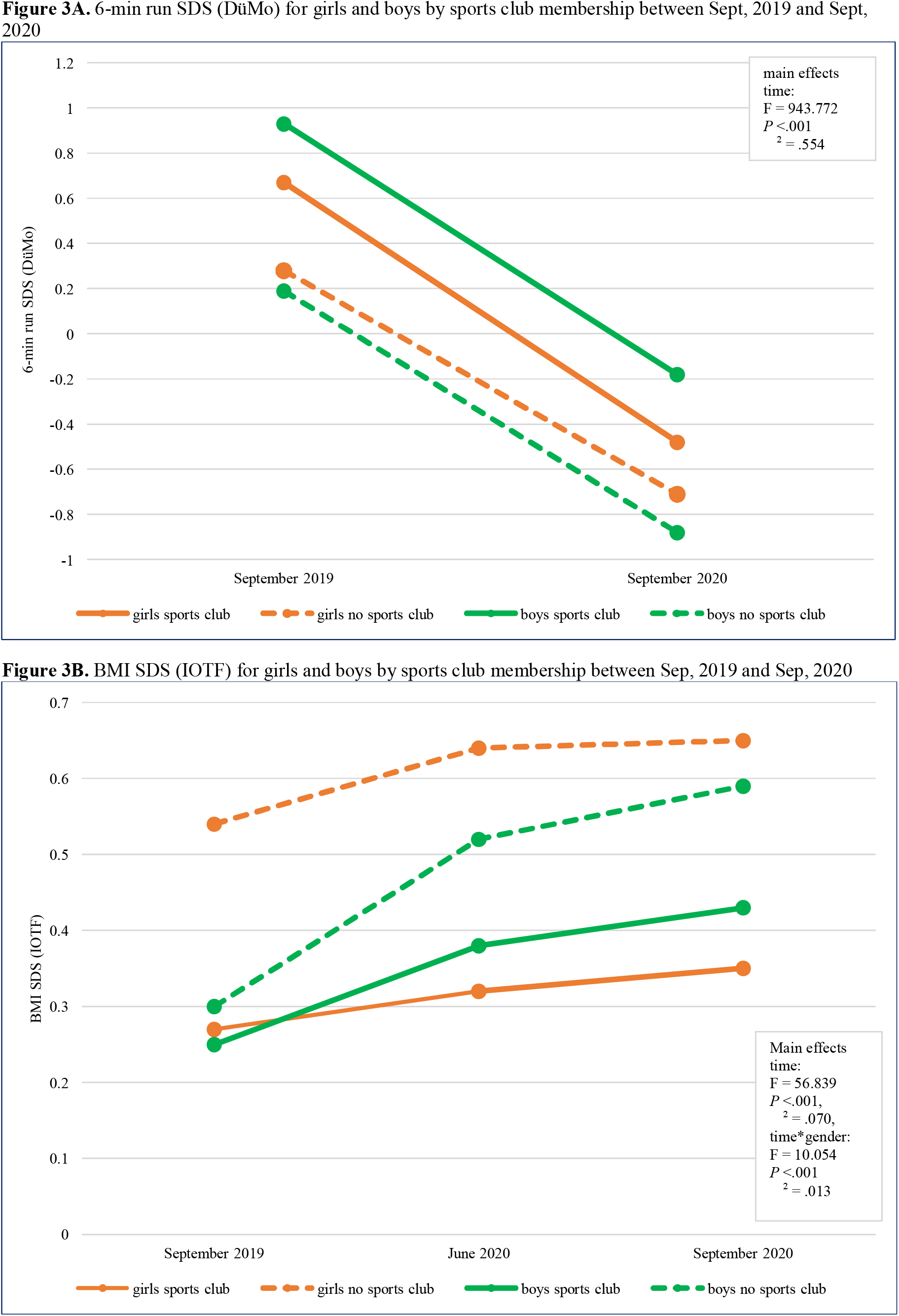
Changes in cardiorespiratory fitness and body mass index between Sept, 2019 and Sept, 2020

### Change in BMI

Due to a reduced energy expenditure related to a reduction in physical activity and sport performance, body fatness might be affected. We observed a significant increase in BMI SDS (IOTF) over time (medium sized main effect: η_p_^2^=0.070, *P*<.001). The increase from September 2019 (M 0.37, SD 1.08) to June 2020 (M 0.49, SD 1.10) was more pronounced than that from June 2020 to September 2020 (M 0.53, SD 1.10). Boys tended to have a larger increase in BMI SDS over time than girls did (time*gender: η_p_^2^=0.013, *P*<.001). Changes over time for IOTF reference values are shown in Figure 3B (eTable 4 and eTable 5). Means and SDs for all subgroups by time point are presented in Table 1 (eTable 2). Similar changes in EQUI BMI (AUT) and BMI SDS (WHO) data were found (eTable 4 and eTable 5). We observed a parallel, although smaller increase in height over time for Austrian and WHO reference values (eTable 6 and eTable 7). Since increased height with unchanged weight would result in a lower BMI, the increase in BMI observed was even more striking considering this small increase in height SDS over time. MANOVA (combination of BMI SDS and height SDS) results revealed a large main effect over time (AUT: η_p_^2^=0.147, *P*<.001; WHO: η_p_^2^=0.138, *P*<.001); eTable 8).

The increase in BMI SDS (IOTF) resulted in an increased proportion of children classified as overweight or obese. In September 2019, 23.8% of girls and 16.8% of boys were overweight or obese according to IOTF thresholds (Table 1, Figure 4A). The proportion of children who were overweight or obese had increased by 1.8% (1.5% girls, 2.1% boys) in June 2020 and by 3.8% (3.1% girls, 4.5% boys) in September 2020 (Table 1, Figure 4B). Although the proportions of children who were overweight and obese were different when using Austrian (eFigure 1 and eFigure 2) or WHO thresholds, the increase from September 2019 to September 2020 was significant for all three methods used to categorise BMI (eTable 9 and eTable 10).

**Figure 4.**
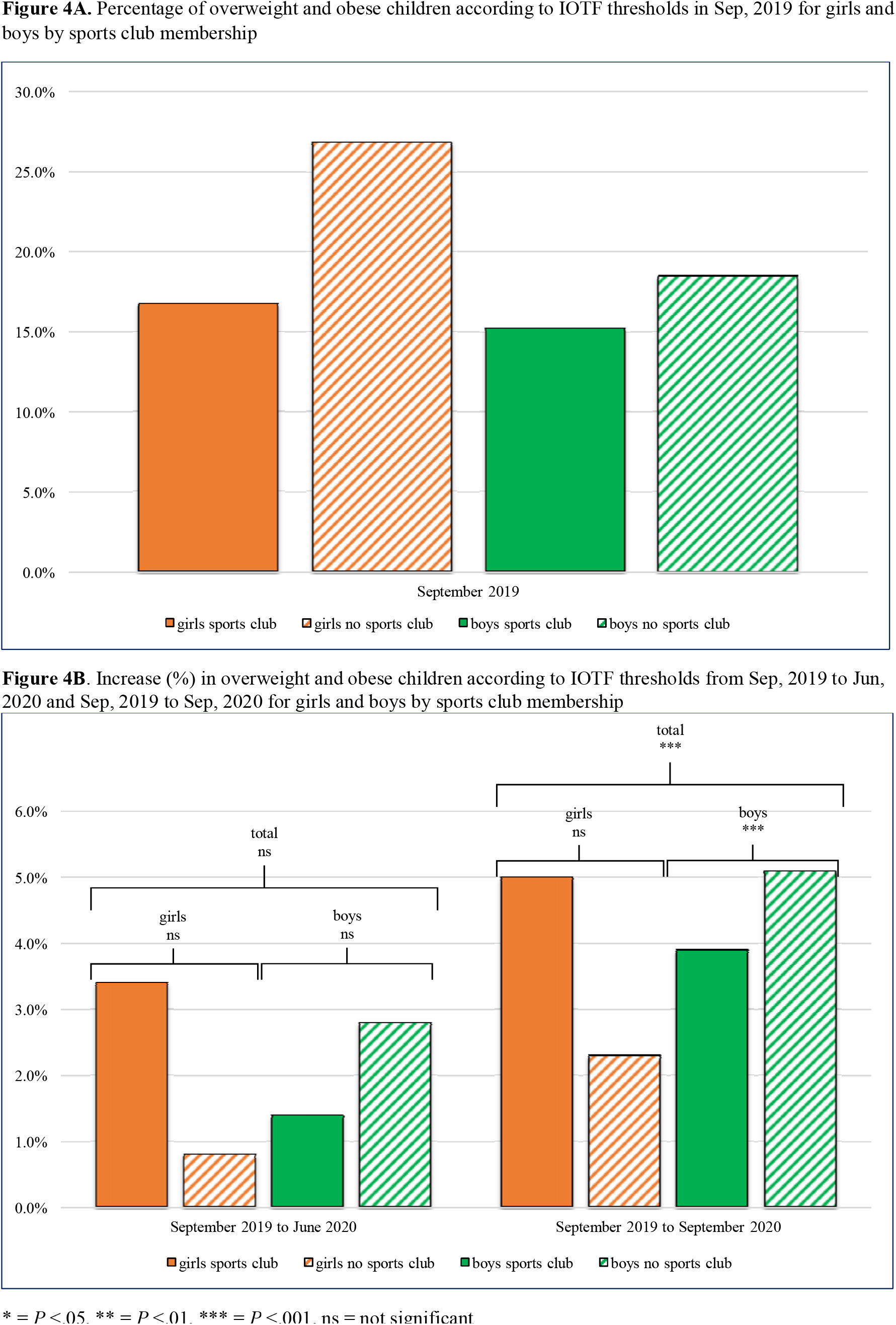
Changes in the proportion of children with overweight or obesity between Sept, 2019 and Sept, 2020

In the subgroup level, different patterns of increase in the proportion of children who were overweight and obese were observed, depending on the used reference thresholds (eTable 9-12).

## DISCUSSION

Our study is the first to address the consequences of COVID-19 mitigation measures on objective health-related parameters in a representative sample of primary school children. We observed a reduction in CRF SDS and an increase in BMI SDS from September 2019 to September 2020.

These worrying findings are in line with the results from a small sample-sized study that reported lower CRF in 10 children during lockdown compared to 10 matched controls from before lockdown in the USA.^19^ A study on 25 adolescent elite soccer players reported a 9% reduction in oxygen consumption on aerobic capacity test after 8 weeks of home confinement^20^, despite participating in a home training programme. A lower CRF may be the direct result of a reduction in physical activity levels,^7-10^ especially a reduction in activities of higher intensity levels, such as those performed in PE classes and during sport activities. The magnitude of the decrease in CRF was about 1 SD in all subgroups, defined by gender or sports club membership, which could be qualified as a very large effect (η_p_^2^=0.55). Although the increase in BMI SDS might also be due to other changes in energy-balance-related behaviour such as diet,^5^ a certain contributing factor is the reduction in energy expenditure due to lower physical activity levels. The proportion of children with overweight or obesity increased by 3.1% in girls and 4.5% in boys. A study from Korea, the only other study with data on the impact of the COVID-19 measures on objectively measured BMI, reported an average increase of 0.22 in BMI SDS,^21^ which is higher than what we observed in Austrian children in the current study. This might be due to different methods of selection of the samples (specific reasons for clinic visitation [Korea] vs random selection of primary school children [Austria]), stricter mitigation measures in Korea,^22^ or because BMI outcomes are difficult to compare between Asian and Caucasian populations.^23,24^

### Strengths and limitations

Our study population is representative of children in the whole region of Klagenfurt, representing those in both rural and the urban areas. Participation rates were high and loss-to-follow-up was very low. Therefore, our results can be generalised to the whole Austria, where identical COVID-19 measures were implemented, and possibly to other countries with similar mitigation measures. The objective, longitudinal measurements of weight, height and CRF were unique, since all previous studies assessing the impact of COVID-19 measures on health parameters of children used self-reported data. The two measurements were exactly a year apart (September 2019 and 2020), ruling out seasonal influences on CRF and BMI values. A limitation of our study is that, for obvious reasons, we did not design the study for the purpose of the analyses presented in this paper. We also did not have a control group of children unaffected by the COVID-19 mitigation measures; hence, we must be careful with causal inference. Due to the lack of a control group, and since both the results of the 6-min run test and BMI measurements in growing children change over time, we assessed whether these changes occurred in our sample, relative to several established age- and gender-specific references. Regardless of which reference values we used,^12,13^ the results of the decrease in CRF remained the same. Results from sensitivity analyses for BMI SDS, using national and international reference values, show similar, significant increases in BMI SDS over time. The only differences were found for BMI categories, when using international^16,17^ or Austrian reference values.^14^

### Implications

Why should we worry about these effects in children? One might argue that these will be transitory, that is, children are flexible and will recover from the restrictions imposed on their lives. However, this is not a given, and might not be true for all children. To our knowledge, there is no information on the recovery of healthy children after a long period of enforced inactivity. The results presented here are the changes that occurred until September 2020, and since then children in Austria were again in partial lockdown, which will likely have additional detrimental effects on their health. Future follow-up measurements of our cohort are planned for March, June and September 2021. However, one thing is a given: we must work hard to ensure that children will indeed recover and get back to an age-adequate level of CRF and BMI. This will mean investing time and effort in PE classes in schools and supporting children to be active in their leisure time. Schools should not only focus on catching up on the deficits in academic learning due to COVID-19 measures, which is unarguably extremely important, but they should also prioritise physical development in children. Even when making an all-out effort of tackling this infectious disease, we should not forget about the enormous ‘slow-motion’ pandemic of non-transmittable diseases related to obesity and lack of physical activity.

### Conclusion

In a representative sample of 7–10-year-old children, CRF levels decreased and BMI SDS increased from September 2019 to September 2020, most likely due to the COVID-19 mitigation measures. It is essential that these negative effects on parameters relevant for long-term health of children are reversed as soon as possible.

## Data Availability

Data collected for the study, including individual participant data will not be made available to other, since subsequent follow-up investigations are in progress. When all follow-up investigations are finished, data might be made available upon reasonable request.

## Author Contributions

All authors had full access to all the data in the study and take responsibility for the integrity of the data and the accuracy of the data analysis.

**Concept and design:** Jarnig, Jaunig, van Poppel.

**Acquisition, analysis, or interpretation of data:** Jarnig, Jaunig, van Poppel.

**Drafting of the manuscript:** Jarnig, Jaunig, van Poppel.

**Critical revision of the manuscript for important intellectual content**: Jarnig, Jaunig, van Poppel.

**Statistical analysis:** Jarnig, Jaunig van Poppel.

**Obtained funding:** Jarnig

**Administrative, technical, or material support:** Jarnig.

**Supervision:** Jarnig, Jaunig van Poppel.

## Additional Contributions

This study (and GJ) was funded by the Austrian Federal Ministry for Arts, Culture, Civil Service and Sport (GZ205.410/0014-II/B/5/2018) and organized by the non-profit association NAMOA – Nachwuchsmodell Austria. We would like to thank all participants and their guardians; the trainers and staff of this study; and the director of education of Carinthia, Robert Klinglmair. We also thank Wolfgang Modritz for the initiation of this study; Rodrigo A. Lima and Peter Hofmann (University of Graz) for the support in the conception phase; and Hannes Wolf for his help to continue the assessments after the COVID-19 lockdown. We would like to express our thanks to the Austrian Working Group on Pediatric Endocrinology and Diabetrics (www.wachstum.at), for providing the calculations for the Austrian reference values for height SDS and EQUI BMI. We also appreciate the flexibility and responsiveness of Christian Günter (Austrian Federal Ministry of Sport) in the everchanging situations of this pandemic.

