## supplementary-material for "Changes in cardiorespiratory fitness and body mass index due to COVID-19 mitigation measures in Austrian children aged 7 to 10 years"

### Supplementary Online Content

**eTable 1.** Restrictions for children in Austria from 31 January 2020 to 16 September 2020 in relation to the OxCGRT stringency index

**eTable 2.** Additional sample characteristics for boys and girls by sports club membership

**eTable 3.** 3-way mixed ANOVAs for cardio respiratory fitness (6-min run) SDS using DüMo and GMT reference values

**eTable 4.** 3-way mixed ANOVAs for BMI SDS using IOTF, Austrian and WHO reference values

**eTable 5.** Post-hoc tests for BMI SDS for the main effect time and time*gender interaction based on the estimated marginal means

**eTable 6.** 3-way mixed ANOVAs for height SDS using Austrian and WHO reference values

**eTable 7.** Post-hoc tests for height SDS for the main effect time and time*gender interaction based on the estimated marginal means

**eTable 8.** Two-factor mixed MANOVAs for BMI SDS and height SDS using Austrian and WHO reference values

**eTable 9.** Cochran's Q Test for dichotomous BMI classification using IOTF, Austrian and WHO reference values

**eTable 10.** Post-hoc tests for dichotomous BMI classification using IOTF, Austrian and WHO reference values

**eTable 11.** Cochran's Q Test for dichotomous BMI classification using IOTF, Austrian and WHO reference values

**eTable 12.** Post-hoc tests for dichotomous BMI classification using IOTF, Austrian and WHO reference values

**eFigure 1**. Percentage of overweight and obese children according to the Austrian thresholds in Sep, 2019

**eFigure 2.** Increase (%) in overweight and obese children according to the Austrian thresholds from Sep, 2019 to Jun, 2020 and Sep, 2019 to Sep, 2020

This supplementary material has been provided by the authors to give readers additional information about their work.

**eTable 1.** Restrictions for children in Austria from 31 January 2020 to 16 September 2020 in relation to the OxCGRT stringency index

| **Period** | **OxCGRT Austria** | **Sschool structure** | **Physical education in school** | **Sports and leisure facilities** | **Sport at the club** | **Stay at home requirements** |
|---|---|---|---|---|---|---|
| 24.02.2020 – 08.03.2020 | 11.11 | 0 | 0 | 0 | 0 | 0 |
| 09.03.2020 – 11.03.2020 | 19.44 | 0 | 0 | 0 | 0 | 0 |
| 12.03.2020 | 34.26 | 0 | 0 | 0 | 0 | 0 |
| 13.03.2020 – 15.03.2020 | 48.15 | 0 | 0 | 0 | 0 | 0 |
| 16.03.2020 – 13.04.2020 | 81.48 | 3 | 3 | 3 | 3 | 3 |
| 14.04.2020 – 22.04.2020 | 77.78 | 3 | 3 | 3 | 3 | 3 |
| 23.04.2020 – 30.04.2020 | 75.00 | 3 | 3 | 3 | 3 | 3 |
| 01.05.2020 | 67.59 | 3 | 3 | 1 | 3 | 1 |
| 02.05.2020 – 10.05.2020 | 64.81 | 3 | 3 | 1 | 3 | 1 |
| 11.05.2020 – 17.05.2020 | 59.26 | 3 | 3 | 1 | 3 | 1 |
| 18.05.2020 – 28.05.2020 | 59.26 | 2 | 3 | 1 | 3 | 1 |
| 29.05.2020 – 02.06.2020 | 53.70 | 2 | 3 | 1 | 2 | 1 |
| 03.06.2020 | 50.00 | 2 | 3 | 1 | 2 | 1 |
| 04.06.2020 – 1.07.2020 | 47.22 | 2 | 3 | 1 | 2 | 1 |
| 02.07.2020 – 09.07.2020 | 47.22 | 2 | 3 | 0 | 0 | 1 |
| 10.07.2020 – 05.09.2020 | 35.19 | 0 | 0 | 0 | 0 | 1 |
| 06.09.2020 – 13.09.2020 | 36.11 | 0 | 0 | 0 | 0 | 1 |
| 14.09.2020 – 16.09.2020 | 36.11 | 0 | 0 | 0 | 0 | 1 |
| 17.09.2020 – 28.09.2020 | 37.04 | 0 | 0 | 0 | 0 | 1 |
| 29.09.2020 – 30.09.2020 | 40.74 | 0 | 0 | 0 | 0 | 1 |

0 = no restrictions or holidays, 1 = low restrictions, 2 = medium restrictions, 3 = high restrictions
OxCGRT = Oxford COVID-19 Government Response Tracker Stringency Index for Austria

**eTable 2.** Additional sample characteristics for boys and girls by sports club membership

| | | | Sep 2019 | Jun 2020 | Sep 2020 |
|---|---|---|---|---|---|
| **Sports Club Membership, No. (%)** | girls | sports club | 119 (15.6%) | 119 (15.6%) | 119 (15.6%) |
| | | no sports club | 264 (34.6%) | 264 (34.6%) | 264 (34.6%) |
| | boys | sports club | 203 (26.6%) | 203 (26.6%) | 203 (26.6%) |
| | | no sports club | 178 (23.3%) | 178 (23.3%) | 178 (23.3%) |
| **6-min run, mean (SD), m** | girls | sports club | 900.9 (113.4) | ND | 797.0 (127.8) |
| | | no sports club | 857.8 (122.8) | ND | 768.6 (113.6) |
| | boys | sports club | 1005.5 (126.7) | ND | 897.3 (126.6) |
| | | no sports club | 915.0 (148.6) | ND | 802.4 (134.6) |
| **6-min run SDS (DüMo), mean (SD)** | girls | sports club | 0.67 (1.00) | ND | - 0.48 (1.02) |
| | | no sports club | 0.28 (1.09) | ND | - 0.71 (0.92) |
| | boys | sports club | 0.93 (1.06) | ND | - 0.18 (0.92) |
| | | no sports club | 0.19 (1.17) | ND | - 0.88 (0.91) |
| **6-min run SDS (GMT), mean (SD)** | total | | 0.15 (1.08) | ND | - 0.96 (1.01) |
| | girls | total | 0.12 (1.03) | ND | - 0.96 (0.98) |
| | | sports club | 0.37 (0.94) | ND | - 0.80 (1.02) |
| | | no sports club | - 0.01 (1.05) | ND | - 1.03 (0.96) |
| | boys | total | 0.18 (1.14) | ND | - 0.96 (1.04) |
| | | sports club | 0.53 (1.00) | ND | - 0.62 (0.94) |
| | | no sports club | - 0.21 (1.15) | ND | - 1.35 (1.01) |
| **Weight, mean (SD), kg** | girls | sports club | 28.7 (6.1) | 31.6 (7.2) | 32.9 (7.8) |
| | | no sports club | 30.4 (7.4) | 33.8 (8.7) | 35.2 (9.3) |
| | boys | sports club | 29.3 (6.1) | 32.4 (7.5 | 33.6 (7.7) |
| | | no sports club | 30.3 (8.5) | 33.9 (9.7 | 35.3 (10.2) |
| **Height, mean (SD), cm** | girls | sports club | 131.3 (6.6) | 135.6 (6.9) | 137.3 (7.1) |
| | | no sports club | 131.9 (6.9) | 136.2 (7.2) | 138.0 (7.2) |
| | boys | sports club | 132.7 (6.3) | 136.7 (6.6) | 138.3 (6.6) |
| | | no sports club | 132.8 (6.5) | 137.0 (6.6) | 138.5 (6.7) |
| **BMI SDS (IOTF), mean (SD)** | girls | sports club | 0.27 (0.96) | 0.32 (1.00) | 0.35 (1.06) |
| | | no sports club | 0.54 (1.13) | 0.64 (1.13) | 0.65 (1.15) |
| | boys | sports club | 0.25 (0.96) | 0.38 (1.02) | 0.43 (0.97) |
| | | no sports club | 0.30 (1.20) | 0.52 (1.18) | 0.59 (1.17) |
| **EQUI BMI (AUT) , mean (SD)** | total | | 22.2 (3.5) | 22.6 (3.8) | 22.7 (3.8) |
| | girls | total | 22.1 (3.5) | 22.3 (3.6) | 22.3 (3.7) |
| | | sports club | 21.4 (2.7) | 21.5 (2.9) | 21.6 (3.1) |
| | | no sports club | 22.4 (3.7) | 22.6 (3.8) | 22.7 (3.9) |
| | boys | total | 22.4 (3.6) | 22.9 (3.9) | 23.0 (4.0) |
| | | sports club | 22.2 (2.8) | 22.5 (3.1) | 22.6 (3.0) |
| | | no sports club | 22.6 (4.3) | 23.3 (4.6) | 23.5 (4.8) |
| **BMI SDS (WHO), mean (SD)** | total | | 0.35 (1.21) | 0.50 (1.24) | 0.55 (1.25) |
| | girls | total | 0.41 (1.16) | 0.50 (1.18) | 0.52 (1.23) |
| | | sports club | 0.22 (1.01) | 0.26 (1.07) | 0.29 (1.14) |
| | | no sports club | 0.50 (1.21) | 0.61 (1.22) | 0.62 (1.25) |
| | boys | total | 0.29 (1.27) | 0.50 (1.30) | 0.57 (1.26) |
| | | sports club | 0.26 (1.12) | 0.41 (1.19) | 0.47 (1.13) |
| | | no sports club | 0.34 (1.43) | 0.60 (1.41) | 0.68 (1.40) |

Sample size for anthropometric data, N = 764; for 6-min run test, N = 763.
EQUI BMI (AUT) = equivalent BMI based on Austrian reference centile curves passing through adult BMI values (Mayer et al, 2015), BMI = body mass index, cm = centimetre, DüMo = 6-min run SDS based on the Düsseldorfer Modell (Stemper et al, 2020), GMT = 6-min run SDS based on the German Motor Test (Bös et al, 2016), IOTF = BMI SDS based on International Obesity Taskforce reference centile curves (Cole et al, 2012), kg = kilogram, m = meter, ND = not determined, SDS = standard deviation score, WHO = BMI SDS based on World Health Organization reference centile curves (de Onis et al, 2007).

**eTable 3.** 3-way mixed ANOVAs for cardio respiratory fitness (6-min run) SDS using DüMo and GMT reference values

| | | **Effects** | **df** | **F** | ***P* Value** | $\eta_p^2$ | **Power**[a] |
|---|---|---|---|---|---|---|---|
| **DüMo (Stemper et al)** | Between-subjects effects | Gender | 1 | 1.385 | .240 | .002 | .217 |
| | | Sports Club | 1 | 58.204 | <.001 | .071 | >.99 |
| | | Gender*Sports Club | 1 | 9.213 | .002 | .012 | .858 |
| | | Error | 759 | | | | |
| | Within-subjects effects | Time (T1-T3) | 1 | 943.772 | <.001 | .554 | >.99 |
| | | Time*Gender | 1 | 0.064 | .801 | <.001 | .057 |
| | | Time*Sports Club | 1 | 1.750 | .186 | .002 | .262 |
| | | Time*Gender*Sports Club | 1 | 0.735 | .392 | .001 | .137 |
| | | Error (Time) | 759 | | | | |
| **GMT (Bös et al)** | Between-subjects effects | Gender | 1 | 0.451 | .502 | .001 | .103 |
| | | Sports Club | 1 | 58.080 | <.001 | .071 | >.99 |
| | | Gender*Sports Club | 1 | 10.150 | .002 | .013 | .889 |
| | | Error | 759 | | | | |
| | Within-subjects effects | Time (T1-T3) | 1 | 1112.765 | <.001 | .595 | >.99 |
| | | Time*Gender | 1 | 0.303 | .582 | <.001 | .085 |
| | | Time*Sports Club | 1 | 1.291 | .256 | .002 | .206 |
| | | Time*Gender*Sports Club | 1 | 0.918 | .338 | .001 | .160 |
| | | Error (Time) | 759 | | | | |

[a] observed power computed using alpha = .05

ANOVA = analysis of variance, df = degrees of freedom, DüMo = 6-min run SDS based on the Düsseldorfer Modell (Stemper et al, 2020), $\eta_p^2$ = partial eta square, GMT = 6-min run SDS based on the German Motor Test (Bös et al, 2016), SDS = standard deviation score.

**eTable 4.** 3-way mixed ANOVAs for BMI SDS using IOTF, Austrian and WHO reference values

| | | **Effects** | **df** | **F** | ***P* Value** | $\eta_p^2$ | **Power[a]** |
|---|---|---|---|---|---|---|---|
| **IOTF (Cole et al)** | Between-subjects effects | Gender | 1 | 0.368 | .544 | <.001 | .093 |
| | | Sports Club | 1 | 6.683 | .010 | .009 | .733 |
| | | Gender*Sports Club | 1 | 1.216 | .271 | .002 | .196 |
| | | Error | 760 | | | | |
| | Within-subjects effects | Time (T1-T2-T3) | 1.960 | 56.839 | <.001 | .070 | >.99 |
| | | Time*Gender | 1.960 | 10.054 | <.001 | .013 | .984 |
| | | Time*Sports Club | 1.960 | 3.924 | .021 | .005 | .702 |
| | | Time*Gender*Sports Club | 1.960 | 0.726 | .482 | .001 | .172 |
| | | Error (Time) | 1489.785 | | | | |
| **AUT (Mayer et al)** | Between-subjects effects | Gender | 1 | 7.215 | .007 | .009 | .765 |
| | | Sports Club | 1 | 10.265 | .001 | .013 | .893 |
| | | Gender*Sports Club | 1 | 0.281 | .596 | <.001 | .083 |
| | | Error | 760 | | | | |
| | Within-subjects effects | Time (T1-T2-T3) | 1.880 | 52.192 | <.001 | .064 | >.99 |
| | | Time*Gender | 1.880 | 12.725 | <.001 | .016 | .996 |
| | | Time*Sports Club | 1.880 | 5.714 | .004 | .007 | .850 |
| | | Time*Gender*Sports Club | 1.880 | 2.319 | .102 | .003 | .456 |
| | | Error (Time) | 1429.154 | | | | |
| **WHO (de Onis et al)** | Between-subjects effects | Gender | 1 | 0.222 | .637 | <.001 | .076 |
| | | Sports Club | 1 | 6.911 | .009 | .009 | .747 |
| | | Gender*Sports Club | 1 | 0.808 | .369 | .001 | .146 |
| | | Error | 760 | | | | |
| | Within-subjects effects | Time (T1-T2-T3) | 1.954 | 60.656 | <.001 | .074 | >.99 |
| | | Time*Gender | 1.954 | 13.701 | <.001 | .018 | .998 |
| | | Time*Sports Club | 1.954 | 3.840 | .023 | .005 | .690 |
| | | Time*Gender*Sports Club | 1.954 | 0.629 | .530 | .001 | .154 |
| | | Error (Time) | 1484.949 | | | | |

[a] observed power computed using alpha = .05

ANOVA = analysis of variance, AUT = equivalent BMI based on Austrian reference centile curves passing through adult BMI values (Mayer et al, 2015), BMI = body mass index, df = degrees of freedom, $\eta_p^2$ = partial eta square, IOTF = BMI SDS based on International Obesity Taskforce reference centile curves (Cole et al, 2012), SDS = standard deviation score, WHO = BMI SDS based on World Health Organization reference centile curves (de Onis et al, 2007).

**eTable 5.** Post-hoc tests for BMI SDS for the main effect time and time*gender interaction based on the estimated marginal means

| | | Pairwise comparisons | Mean diff (95% CI) | SE | p-lvl | *P* Value[a] |
|---|---|---|---|---|---|---|
| **IOTF (Cole et al)** | Time | T1 vs T2 | -0.124 (-0.163 to -0.085) | 0.016 | *** | <.001 |
| | | T1 vs T3 | -0.163 (-0.203 to -0.123) | 0.017 | *** | <.001 |
| | | T2 vs T3 | -0.039 (-0.074 to -0.003) | 0.015 | * | .026 |
| | Time*Gender (Girls) | T1 vs T2 | -0.075 (-0.133 to -0.018) | 0.024 | ** | .005 |
| | | T1 vs T3 | -0.093 (-0.152 to -0.035) | 0.024 | *** | <.001 |
| | | T2 vs T3 | -0.018 (-0.070 to 0.034) | 0.022 | | 1.000 |
| | Time*Gender (Boys) | T1 vs T2 | -0.173 (-0.226 to -0.119) | 0.022 | *** | <.001 |
| | | T1 vs T3 | -0.233 (-0.287 to -0.178) | 0.023 | *** | <.001 |
| | | T2 vs T3 | -0.060 (-0.108 to -0.012) | 0.020 | ** | .009 |
| **AUT (Mayer et al)** | Time | T1 vs T2 | -0.307 (-0.413 to -0.201) | 0.044 | *** | <.001 |
| | | T1 vs T3 | -0.428 (-0.541 to -0.314) | 0.047 | *** | <.001 |
| | | T2 vs T3 | -0.121 (-0.211 to -0.030) | 0.038 | ** | .004 |
| | Time*Gender (Girls) | T1 vs T2 | -0.141 (-0.296 to 0.014) | 0.065 | | .089 |
| | | T1 vs T3 | -0.223 (-0.388 to -0.057) | 0.069 | ** | .004 |
| | | T2 vs T3 | -0.082 (-0.215 to 0.050) | 0.055 | | .412 |
| | Time*Gender (Boys) | T1 vs T2 | -0.473 (-0.617 to -0.329) | 0.060 | *** | <.001 |
| | | T1 vs T3 | -0.632 (-0.786 to -0.478) | 0.064 | *** | <.001 |
| | | T2 vs T3 | -0.159 (-0.283 to -0.036) | 0.051 | ** | .006 |
| **WHO (de Onis et al)** | Time | T1 vs T2 | -0.143 (-0.187 to -0.099) | 0.018 | *** | <.001 |
| | | T1 vs T3 | -0.190 (-0.235 to -0.144) | 0.019 | *** | <.001 |
| | | T2 vs T3 | -0.047 (-0.087 to -0.007) | 0.019 | * | .014 |
| | Time*Gender (Girls) | T1 vs T2 | -0.076 (-0.141 to -0.012) | 0.027 | * | .013 |
| | | T1 vs T3 | -0.099 (-0.165 to -0.033) | 0.028 | *** | <.001 |
| | | T2 vs T3 | -0.022 (-0.081 to 0.036) | 0.024 | | 1.000 |
| | Time*Gender (Boys) | T1 vs T2 | -0.209 (-0.269 to -0.149) | 0.025 | *** | <.001 |
| | | T1 vs T3 | -0.280 (-0.342 to -0.219) | 0.026 | *** | <.001 |
| | | T2 vs T3 | -0.071 (-0.125 to -0.017) | 0.023 | ** | .005 |

[a] adjusted for multiple comparisons using Bonferroni correction.

p-lvl (*P* Value level) * = *P* <.05, * = *P* <.01, * = *P* <.001, BMI = body mass index, CI = confidence interval, AUT = equivalent BMI based on Austrian reference centile curves passing through adult BMI values (Mayer et al, 2015), IOTF = BMI SDS based on International Obesity Taskforce reference centile curves (Cole et al, 2012), Mean diff = mean difference based on the estimated marginal means, p-lvl = significance level, SDS = standard deviation score, SE = standard error, T1= baseline measurements in Sept and Oct 2019, T2 = follow-up measurements in May and Jun 2020, T2 = follow-up measurements in Sep and Oct 2020, WHO = BMI SDS based on World Health Organization reference centile curves (de Onis et al, 2007).

**eTable 6.** 3-way mixed ANOVAs for height SDS using Austrian and WHO reference values

| | | **Effects** | **df** | **F** | ***P* Value** | $\eta_p^2$ | **Power**[a] |
|---|---|---|---|---|---|---|---|
| **AUT (Gleiss et al)** | Between-subjects effects | Gender | 1 | 1.354 | .245 | .002 | .213 |
| | | Sports Club | 1 | 0.058 | .810 | <.001 | .057 |
| | | Gender*Sports Club | 1 | 1.417 | .234 | .002 | .221 |
| | | Error | 760 | | | | |
| | Within-subjects effects | Time (T1-T2-T3) | 1.799 | 25.730 | <.001 | .033 | >.99 |
| | | Time*Gender | 1.799 | 5.473 | .006 | .007 | .821 |
| | | Time*Sports Club | 1.799 | 1.293 | .273 | .002 | .267 |
| | | Time*Gender*Sports Club | 1.799 | 1.889 | .156 | .002 | .373 |
| | | Error (Time) | 1366.918 | | | | |
| **WHO (de Onis et al)** | Between-subjects effects | Gender | 1 | 2.247 | .134 | .003 | .322 |
| | | Sports Club | 1 | 0.050 | .823 | <.001 | .056 |
| | | Gender*Sports Club | 1 | 1.322 | .251 | .002 | .209 |
| | | Error | 760 | | | | |
| | Within-subjects effects | Time (T1-T2-T3) | 1.791 | 7.939 | .001 | .010 | .938 |
| | | Time*Gender | 1.791 | 7.395 | .001 | .010 | .921 |
| | | Time*Sports Club | 1.791 | 1.597 | .205 | .002 | .321 |
| | | Time*Gender*Sports Club | 1.791 | 1.440 | .238 | .002 | .293 |
| | | Error (Time) | 1360.842 | | | | |

[a] observed power computed using alpha = .05

ANOVA = analysis of variance, AUT = height SDS based on Austrian reference centile curves (Gleiss et al, 2013), df = degrees of freedom, $\eta_p^2$ = partial eta square, SDS = standard deviation score, WHO = height SDS based on World Health Organization reference centile curves (de Onis et al, 2007).

**eTable 7.** Post-hoc tests for height SDS for the main effect time and time*gender interaction based on the estimated marginal means

| | | Pairwise comparison | Mean diff (95% CI) | SE | p-lvl | *P* Value[a] |
|---|---|---|---|---|---|---|
| **AUT (Gleiss et al)** | Time | T1 vs T2 | -0.040 (-0.055 to -0.025) | 0.006 | *** | <.001 |
| | | T1 vs T3 | -0.039 (-0.056 to -0.021) | 0.007 | *** | <.001 |
| | | T2 vs T3 | 0.001 (-0.011 to 0.014) | 0.005 | | >.99 |
| | Time*Gender (Girls) | T1 vs T2 | -0.052 (-0.074 to -0.029) | 0.009 | *** | <.001 |
| | | T1 vs T3 | -0.060 (-0.085 to -0.034) | 0.010 | *** | <.001 |
| | | T2 vs T3 | -0.008 (-0.026 to 0.011) | 0.008 | | .951 |
| | Time*Gender (Boys) | T1 vs T2 | -0.028 (-0.049 to -0.007) | 0.009 | ** | .004 |
| | | T1 vs T3 | -0.018 (-0.041 to 0.006) | 0.010 | | .208 |
| | | T2 vs T3 | 0.011 (-0.007 to 0.028) | 0.007 | | .431 |
| **WHO (de Onis et al)** | Time | T1 vs T2 | -0.021 (-0.035 to -0.006) | 0.006 | ** | .002 |
| | | T1 vs T3 | -0.021 (-0.037 to -0.004) | 0.007 | ** | .007 |
| | | T2 vs T3 | -0.001 (-0.012 to 0.012) | 0.005 | | >.99 |
| | Time*Gender (Girls) | T1 vs T2 | -0.003 (-0.024 to 0.019) | 0.009 | | >.99 |
| | | T1 vs T3 | 0.001 (-0.023 to 0.025) | 0.010 | | >.99 |
| | | T2 vs T3 | 0.003 (-0.014 to 0.021) | 0.007 | | >.99 |
| | Time*Gender (Boys) | T1 vs T2 | -0.039 (-0.059 to -0.019) | 0.008 | *** | <.001 |
| | | T1 vs T3 | -0.042 (-0.065 to -0.020) | 0.009 | *** | <.001 |
| | | T2 vs T3 | -0.004 (-0.020 to 0.013) | 0.007 | | >.99 |

[a] adjusted for multiple comparisons using Bonferroni correction.
p-lvl (*P* Value level) * = *P* <.05, * = *P* <.01, * = *P* <.001, CI = confidence interval, AUT = height SDS based on Austrian reference centile curves (Gleiss et al, 2013), Mean diff = mean difference based on the estimated marginal means, p-lvl = significance level, SDS = standard deviation score, SE = standard error, T1= baseline measurements in Sept and Oct 2019, T2 = follow-up measurements in May and Jun 2020, T3 = follow-up measurements in Sep and Oct 2020, WHO = height SDS based on World Health Organization reference centile curves (de Onis et al, 2007).

**eTable 8.** Two-factor mixed MANOVAs for BMI SDS and height SDS using Austrian and WHO reference values

| | | **Λ** | **df** | **F** | ***P* Value** | $\boldsymbol{\eta_p^2}$ | **Power**[a] |
|---|---|---|---|---|---|---|---|
| **AUT (Mayer et al)** | Time | 0.853 | 757 | 32.590 | <.001 | .147 | >.99 |
| | Time*Gender | 0.965 | 757 | 6.834 | <.001 | .035 | .994 |
| | Time*Sports club | 0.985 | 757 | 2.979 | .019 | .015 | .796 |
| | Time*Gender*Sports club | 0.989 | 757 | 2.190 | .068 | .011 | .647 |
| **WHO (de Onis et al)** | Time | 0.862 | 757 | 30.325 | <.001 | .138 | >.99 |
| | Time*Gender | 0.954 | 757 | 9.148 | <.001 | .046 | >.99 |
| | Time*Sports club | 0.987 | 757 | 2.512 | .041 | .013 | .715 |
| | Time*Gender*Sports club | 0.993 | 757 | 1.342 | .253 | .007 | .421 |

[a] observed power computed using alpha = .05

AUT = equivalent BMI based on Austrian reference centile curves passing through adult BMI values (Mayer et al, 2015) and height SDS based on Austrian reference centile curves (Gleiss et al, 2013), BMI = body mass index, df = degrees of freedom, MANOVA = multivariate analysis of variance, $\eta_p^2$ = partial eta square, **Λ** = Wilks' Lambda, SDS = standard deviation score, WHO = BMI SDS and height SDS based on World Health Organization reference centile curves (de Onis et al, 2007).

**eTable 9.** Cochran's Q Test for dichotomous BMI classification using IOTF, Austrian and WHO reference values

| | | n | Q | df | p-lvl | *P* Value |
|---|---|---|---|---|---|---|
| **IOTF (Cole et al)** | total | 764 | 18.029 | 2 | *** | <.001 |
| | girls | 383 | 5.143 | 2 | | .076 |
| | boys | 381 | 15.500 | 2 | *** | <.001 |
| **AUT (Mayer et al)** | total | 764 | 27.360 | 2 | *** | <.001 |
| | girls | 383 | 13.364 | 2 | ** | .001 |
| | boys | 381 | 15.643 | 2 | *** | <.001 |
| **WHO (de Onis et al)** | total | 764 | 29.216 | 2 | *** | <.001 |
| | girls | 383 | 3.526 | 2 | | .172 |
| | boys | 381 | 31.051 | 2 | *** | <.001 |

p-lvl (*P* Value level)  * = *P* <.05, * = *P* <.01, * = *P* <.001, AUT = Austrian reference values using centile curves for BMI 25.00 at the age of 18 as thresholds for overweight and above (Mayer et al, 2015), BMI = body mass index, Q = Cochran`s Q test statistic, df = degree of freedom, IOTF = International Obesity Taskforce reference values using centile curves for BMI 25.00 at the age of 18 as thresholds for overweight and above (Cole et al, 2012), p-lvl = significance level, SDS = standard deviation score, WHO = World Health Organization reference values using +1 SDS percentiles as thresholds for overweight and above (de Onis et al, 2007).

**eTable 10.** Post-hoc tests for dichotomous BMI classification using IOTF, Austrian and WHO reference values

| | | Pairwise comparison | STS | SE | p-lvl | *P* Value[b] |
|---|---|---|---|---|---|---|
| **IOTF (Cole et al)** | total | T1 vs T2 | 2.049 | 0.009 | | .121 |
| | | T1 vs T3 | 4.245 | 0.009 | *** | <.001 |
| | | T2 vs T3 | 2.196 | 0.009 | | .084 |
| | girls | T1 vs T2[a] | | | | |
| | | T1 vs T3 [a] | | | | |
| | | T2 vs T3 [a] | | | | |
| | boys | T1 vs T2 | 1.852 | 0.011 | | .192 |
| | | T1 vs T3 | 3.935 | 0.011 | *** | <.001 |
| | | T2 vs T3 | 2.083 | 0.011 | | .112 |
| **AUT (Mayer et al)** | total | T1 vs T2 | 3.118 | 0.008 | ** | .005 |
| | | T1 vs T3 | 5.196 | 0.008 | *** | <.001 |
| | | T2 vs T3 | 2.078 | 0.008 | | .113 |
| | girls | T1 vs T2 | 2.872 | 0.010 | * | .012 |
| | | T1 vs T3 | 3.395 | 0.010 | ** | .002 |
| | | T2 vs T3 | 0.522 | 0.010 | | >.99 |
| | boys | T1 vs T2 | 1.620 | 0.011 | | .316 |
| | | T1 vs T3 | 3.935 | 0.011 | *** | <.001 |
| | | T2 vs T3 | 2.315 | 0.011 | | .062 |
| **WHO (de Onis et al)** | total | T1 vs T2 | 4.104 | 0.011 | *** | <.001 |
| | | T1 vs T3 | 5.099 | 0.011 | *** | <.001 |
| | | T2 vs T3 | 0.995 | 0.011 | | .959 |
| | girls | T1 vs T2 [a] | | | | |
| | | T1 vs T3 [a] | | | | |
| | | T2 vs T3 [a] | | | | |
| | boys | T1 vs T2 | 3.827 | 0.016 | *** | <.001 |
| | | T1 vs T3 | 5.421 | 0.016 | *** | <.001 |
| | | T2 vs T3 | 1.594 | 0.016 | | .332 |

[a] pairwise comparisons were not performed due to non-significant omnibus test.
[b] adjusted for multiple comparisons using Bonferroni correction.
p-lvl (*P* Value level) * = *P* <.05, * = *P* <.01, * = *P* <.001, AUT = Austrian reference values using centile curves for BMI 25.00 at the age of 18 as thresholds for overweight and above (Mayer et al, 2015), BMI = body mass index, IOTF = International Obesity Taskforce reference values using centile curves for BMI 25.00 at the age of 18 as thresholds for overweight and above (Cole et al, 2012), p-lvl = significance level, SDS = standard deviation score, SE = standard error, STS = standard test statistic, T1= baseline measurements in Sept and Oct 2019, T2 = follow-up measurements in May and Jun 2020, T3 = follow-up measurements in Sep and Oct 2020, WHO = World Health Organization reference values using +1 SDS percentiles as thresholds for overweight and above (de Onis et al, 2007).

**eTable 11.** Cochran's Q Test for dichotomous BMI classification using IOTF, Austrian and WHO reference values

| | | | n | Q | df | p-lvl | *P* Value |
|---|---|---|---|---|---|---|---|
| **IOTF (Cole et al)** | girls | sports club | 119 | 3.733 | 2 | | .155 |
| | | no sports club | 264 | 2.074 | 2 | | .355 |
| | boys | sports club | 203 | 6.533 | 2 | * | .038 |
| | | no sports club | 178 | 9.385 | 2 | ** | .009 |
| **AUT (Mayer et al)** | girls | sports club | 119 | 7.714 | 2 | * | .021 |
| | | no sports club | 264 | 7.600 | 2 | * | .022 |
| | boys | sports club | 203 | 4.933 | 2 | | .085 |
| | | no sports club | 178 | 11.692 | 2 | ** | .003 |
| **WHO (de Onis et al)** | girls | sports club | 119 | 1.273 | 2 | | .529 |
| | | no sports club | 264 | 2.741 | 2 | | .254 |
| | boys | sports club | 203 | 9.484 | 2 | ** | .009 |
| | | no sports club | 178 | 24.071 | 2 | *** | <.001 |

p-lvl (*P* Value level) * = *P* <.05, * = *P* <.01, * = *P* <.001, AUT = Austrian reference values using centile curves for BMI 25.00 at the age of 18 as thresholds for overweight and above (Mayer et al, 2015), BMI = body mass index, Q = Cochran`s Q test statistic, df = degree of freedom, IOTF = International Obesity Taskforce reference values using centile curves for BMI 25.00 at the age of 18 as thresholds for overweight and above (Cole et al, 2012), p-lvl = significance level, SDS = standard deviation score, WHO = World Health Organization reference values using +1 SDS percentiles as thresholds for overweight and above (de Onis et al, 2007).

**eTable 12.** Post-hoc tests for dichotomous BMI classification using IOTF, Austrian and WHO reference values

| | | | **Pairwise comparison** | **STS** | **SE** | **p-lvl** | ***P* Value[b]** |
|---|---|---|---|---|---|---|---|
| **IOTF (Cole et al)** | girls | sports club | T1 vs T2 [a] | | | | |
| | | | T1 vs T3 [a] | | | | |
| | | | T2 vs T3 [a] | | | | |
| | | no sports club | T1 vs T2 [a] | | | | |
| | | | T1 vs T3 [a] | | | | |
| | | | T2 vs T3 [a] | | | | |
| | boys | sports club | T1 vs T2 | -0.949 | 0.016 | | >.99 |
| | | | T1 vs T3 | -2.530 | 0.016 | * | .034 |
| | | | T2 vs T3 | -1.581 | 0.016 | | .342 |
| | | no sports club | T1 vs T2 | -1.698 | 0.017 | | .268 |
| | | | T1 vs T3 | -3.057 | 0.017 | ** | .007 |
| | | | T2 vs T3 | -1.359 | 0.017 | | .523 |
| **AUT (Mayer et al)** | girls | sports club | T1 vs T2 | 1.389 | 0.018 | | .495 |
| | | | T1 vs T3 | 2.777 | 0.018 | * | .016 |
| | | | T2 vs T3 | 1.389 | 0.018 | | .495 |
| | | no sports club | T1 vs T2 | 2.530 | 0.012 | * | .034 |
| | | | T1 vs T3 | 2.214 | 0.012 | | .081 |
| | | | T2 vs T3 | -0.316 | 0.012 | | >.99 |
| | boys | sports club | T1 vs T2 [a] | | | | |
| | | | T1 vs T3 [a] | | | | |
| | | | T2 vs T3 [a] | | | | |
| | | no sports club | T1 vs T2 | -1.359 | 0.017 | | .523 |
| | | | T1 vs T3 | -3.397 | 0.017 | ** | .002 |
| | | | T2 vs T3 | -2.038 | 0.017 | | .125 |
| **WHO (de Onis et al)** | girls | sports club | T1 vs T2 [a] | | | | |
| | | | T1 vs T3 [a] | | | | |
| | | | T2 vs T3 [a] | | | | |
| | | no sports club | T1 vs T2 [a] | | | | |
| | | | T1 vs T3 [a] | | | | |
| | | | T2 vs T3 [a] | | | | |
| | boys | sports club | T1 vs T2 | -2.420 | 0.022 | * | .047 |
| | | | T1 vs T3 | -2.860 | 0.022 | * | .013 |
| | | | T2 vs T3 | -0.440 | 0.022 | | >.99 |
| | | no sports club | T1 vs T2 | -3.009 | 0.024 | ** | .008 |
| | | | T1 vs T3 | -4.861 | 0.024 | *** | <.001 |
| | | | T2 vs T3 | -1.852 | 0.024 | | .192 |

[a] pairwise comparisons were not performed due to non-significant omnibus test.
[b] adjusted for multiple comparisons using Bonferroni correction.
p-lvl (*P* Value level) * = *P* <.05, * = *P* <.01, * = *P* <.001, AUT = Austrian reference values using centile curves for BMI 25.00 at the age of 18 as thresholds for overweight and above (Mayer et al, 2015), BMI = body mass index, IOTF = International Obesity Taskforce reference values using centile curves for BMI 25.00 at the age of 18 as thresholds for overweight and above (Cole et al, 2012), p-lvl = significance level, SDS = standard deviation score, SE = standard error, STS = standard test statistic, T1= baseline measurements in Sept and Oct 2019, T2 = follow-up measurements in May and Jun 2020, T3 = follow-up measurements in Sep and Oct 2020, WHO = World Health Organization reference values using +1 SDS percentiles as thresholds for overweight and above (de Onis et al, 2007).

**eFigure 1**. Percentage of overweight and obese children according to the Austrian thresholds in Sep, 2019

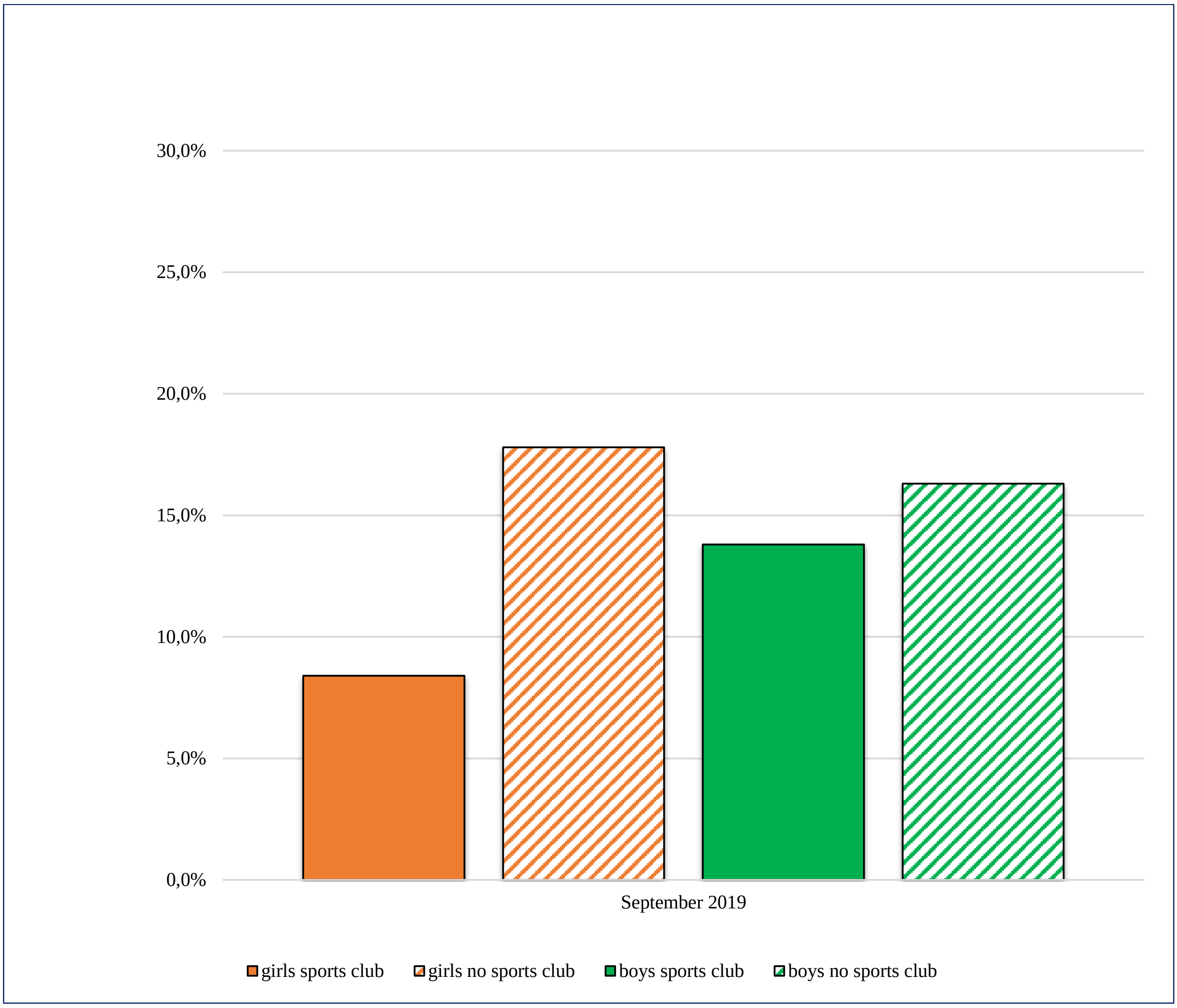

**eFigure 2.** Increase (%) in overweight and obese children according to the Austrian thresholds from Sep, 2019 to Jun, 2020 and Sep, 2019 to Sep, 2020

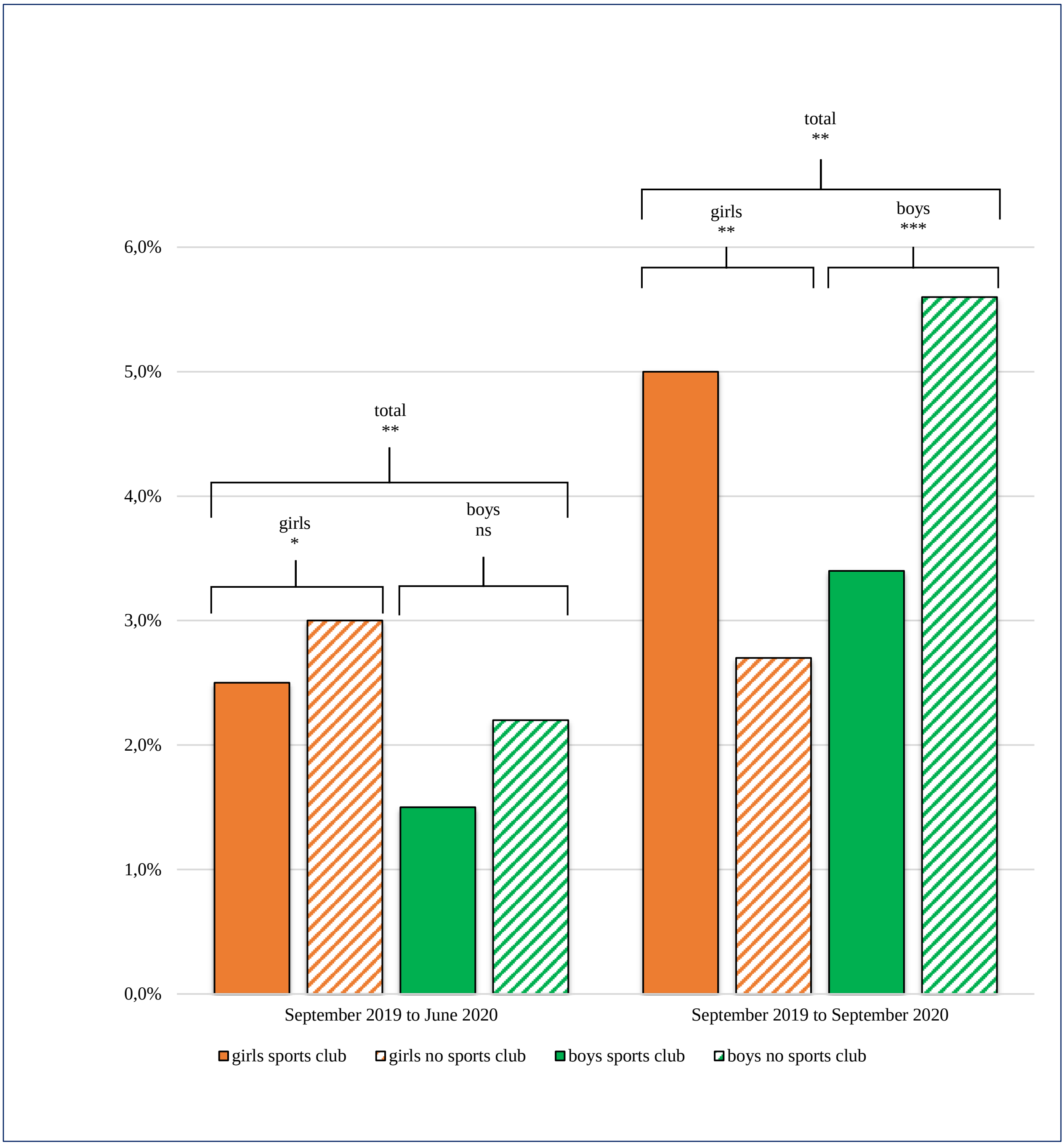


*P* Value level means * = *P* <.05, ** = *P* <.01, *** = *P* <.001, ns = not significant
